# Benchmarking large language models on the Ukrainian Krok 2 licensing examination in physical therapy: accuracy, consistency, and agreement

**DOI:** 10.64898/2026.09.21.26363368

**Authors:** Y.F. Filak, Y.O. Mykhalko, M.V. Sabadosh

## Abstract

**Aim:** To evaluate the accuracy, run-to-run consistency, and agreement of large language models on the Ukrainian Integrated Licensing Examination Krok 2 for the Specialty of Physical Therapy.

**Materials and Methods:** Five cloud-based LLMs (ChatGPT-5.5, Claude 5 Sonnet, Gemini 3.5 Flash, Grok 4, DeepSeek-V3) were evaluated using 150 Ukrainian-language multiple-choice questions from the official Krok 2 database. Each model completed two independent runs with identical inputs. Accuracy (with 95% confidence intervals, Wilson method), run-to-run consistency, and agreement were assessed. Statistical analysis included Cohen’s κ, Cochran’s Q and pairwise McNemar tests with Holm-Bonferroni correction.

**Results:** All models achieved >80% accuracy in both runs, exceeding the passing threshold of ≥64%. In the first run, accuracy ranged from 80.00% (DeepSeek-V3) to 93.33% (Gemini 3.5 Flash); in the second run, from 82.67% (Claude 5 Sonnet) to 88.67% (Gemini 3.5 Flash). In the first run significant differences were observed between Gemini 3.5 Flash and Grok 4 (93.33% vs 84.00%; p=0.0022) and DeepSeek-V3 (93.33% vs 80.00%; p=0.0001). There were no statistically significant differences between models in the second run. R2R consistency exceeded 90% for all models with the highest in ChatGPT-5.5 (98.67%) and lowest in DeepSeek-V3 (90.00%). Cohen’s κ ranged from 0.64 to 0.94, indicating substantial to almost perfect agreement. The stability of LLMs’ response correctness across two runs ranged from 77.33% (DeepSeek-V3) to 88.00% (Gemini 3.5 Flash).

**Conclusions:** All LLMs demonstrated high performance on Ukrainian-language Krok 2 examination tasks. Gemini 3.5 Flash showed the highest accuracy in both runs and response correctness stability while ChatGPT-5.5 exhibited the greatest response consistency and agreement. Although the accuracy and stability of LLM responses may be interrelated, they represent distinct aspects of model performance. This highlights the need for a comprehensive approach to evaluating LLMs. The findings indicate the potential of LLMs as an auxiliary tool in the professional practice of physical therapy specialists.

## Introduction

For the last few years, there has been a rapid development of artificial intelligence (AI) technologies in general and large language models (LLMs) in particular. Their emergence has significantly transformed various industries, including the healthcare sector, where they are considered a promising tool for improving the efficiency of clinical practice, automating work with medical documentation, communicating with patients and conducting scientific research, etc. [1; 2]. In addition, LLMs are actively used in medical education, in particular for creating educational materials, generating clinical scenarios, forming test tasks and providing personalized learning [3]. The application of LLMs in physical therapy and rehabilitation is a relatively new but rapidly growing area of research. Modern models are able to identify functional problems of patients, form rehabilitation appointments and offer reasonable approaches to treatment [4; 5]. In particular, in sports physical therapy, the responses of models can be not only comparable, but also exceed the quality of the responses of junior specialists [6].

There are also wide possibilities for using LLMs to optimize the management of rehabilitation processes and increase the efficiency of health care systems [7]. An important direction is the use of LLMs to create individualized physical therapy programs. Studies show that rehabilitation programs generated by models have a significant level of consistency with expert recommendations. At the same time, they may be inferior in the detail of individual parameters (for example, intensity or progression of exercises), which requires control by specialists [8].

Despite the high potential, the use of LLMs in medicine in general and physical therapy in particular is accompanied by a number of challenges associated with the risk of generating inaccurate or incomplete information, the presence of bias in responses, confidentiality, ethical use and a number of other limitations [9; 10]. At the same time, these technologies are constantly evolving, improving their performance in solving problems in various fields. This creates the necessity for systematic assessment of the quality, reliability and safety of the use of LLMs, as well as the careful development and validation of standardized approaches to this [2]. One of the most common approaches to assessing the capabilities of LLMs is the use of standardized licensing and certification exams, which allows for an objective comparison of their performance with the level of training of medical professionals [11]. A similar approach is used not only in medicine, but also in other subject areas. For example, in programming, standardized problem sets are used, which allows quantitatively comparing the ability of models to think algorithmically, generate correct code, and solve problems of different complexity levels [12].

Modern LLMs have demonstrated high levels of performance on medical-related test tasks. For example, one of the first models in the ChatGPT family has already reached or approached the passing score on the United States Medical Licensing Examination (USMLE) without specialized training [11; 13]. More recent models, such as GPT-4, have demonstrated even higher results, exceeding the passing score and significantly outperforming predecessors [14].

However, most such studies have focused primarily on accuracy measures, while reliability, consistency of results, and clinical relevance are often neglected [15]. However, even high accuracy measures can be misleading without considering the stability of responses and their reproducibility [16].

Furthermore, despite the large number of studies evaluating LLMs on general medical exams, evidence on their effectiveness in highly specialized fields, such as physical therapy and rehabilitation, remains limited.

Another important consideration that may affect both the accuracy and stability of responses is the language in which the query is formulated or the input data is presented. In particular, in a study on diagnosing diseases based on clinical case descriptions, the accuracy of LLMs ranged from 72% for English to 44% for Arabic [17]. Similarly, studies using medical test items have observed statistically significant variations in accuracy ranging from approximately 64% to 87% depending on the language context [18]. In addition, the performance of LLMs may depend on the availability of high-quality data for a specific language, its linguistic features, translation of terms, inaccuracies in wording, and cultural context that affects the interpretation of medical information [17; 19]. There is a complex interaction between the language of the training data, the structure of the query, and the ability of the model to generalize knowledge, and multilingualism issues remain one of the key challenges in the application of LLMs, especially for languages with limited resources [18].

In this context, it is particularly relevant to evaluate the effectiveness of LLMs on tasks formulated in a non-English language, in particular Ukrainian. Such studies allow not only to determine the real level of competence of models in local educational and clinical settings, but also to identify potential risks of their use in practice, where the accuracy and correctness of information interpretation are of critical importance.

## Aim

To evaluate the accuracy, run-to-run consistency, and agreement of large language models on the Ukrainian integrated licensing examination Krok 2 for the specialty of Physical Therapy.

### Object, materials and research methods

The object of the study was the performance of cloud-based LLMs in solving Ukrainian integrated examination Krok 2 multiple-choice questions in the specialty of Physical Therapy.

This research was conducted in July 2025. Five cloud-based LLMs – ChatGPT-5.5 (OpenAI), Claude 5 Sonnet (Anthropic), Gemini 3.5 Flash (Google), Grok 4 (xAI), and DeepSeek-V3 (DeepSeek) – were evaluated to assess their effectiveness in solving test tasks from the Ukrainian integrated licensing examination Krok 2 for the specialty Physical Therapy. Therefore, 150 publicly-available test tasks were selected from the official database [20] and provided to the models. Each model received an identical set of tasks. All questions were in Ukrainian, contained no images, and were formatted as multiple-choice questions (MCQs) with a single best answer. LLMs were used in standard mode without special activation of reasoning functions, deep research modes, external tools, or any other advanced capabilities. The Internet search function, memory function and personalization settings were disabled. The questions were presented sequentially during one session, 10 questions per prompt. This batch size was chosen to balance context length and response stability. All responses were generated within the framework of the user’s normal text interaction with the model via web interface. Only exact matches with the official answer key were considered correct. To assess run-to-run response consistency and agreement, each model was given an identical set of MCQs in two different runs. The order of questions was identical across all models and runs. To avoid the influence of the previous context, a separate chat session was created for each run. At the beginning of each chat, the model received an initial prompt: “I will give you test tasks with answer options for each of them. From the proposed options, choose the most correct one. Write only the task number and the correct answer in the column. Write the correct answer exactly as it is written in the option”. No other prompt engineering techniques were used. The only exception was the initial instruction for the Gemini 3.5 Flash and Grok 4 models – the sentence “Do not look for the answer on the Internet” was added to the initial prompt mentioned above. This was done because these models do not have an option to disable the Internet search function in the settings. The default sampling parameters (temperature, top_p, seed, etc.) specific to each platform were used during all runs as fixed control was not available across all platforms.

## Data processing

All statistical analyses were performed using Python 3.13.7 with the pandas 2.3.1, NumPy 2.3.2, SciPy 1.16.1, statsmodels 0.14.5, and scikit-learn 1.7.1 libraries. Models’ accuracy was defined as the proportion of correct answers with 95% confidence intervals (CIs) calculated using the Wilson method. In this paper, we define run-to-run (R2R) consistency as the proportion of identical model outputs obtained across two independent runs using the same input set.

Cochran’s Q test for related binary samples was used to assess the differences in models’ responses. If Cochran’s Q test showed p-values < 0.05, post hoc pairwise McNemar tests with Yates’ continuity correction and correction for multiple comparisons using the Holm-Bonferroni method were performed. Adjusted p-values < 0.05 were considered statistically significant. To assess the agreement of models within two runs, Cohen’s kappa (κ) was calculated and interpreted according to the following scale: <0.00, poor; 0.00-0.20, slight; 0.21-0.40, fair; 0.41-0.60, moderate; 0.61-0.80, substantial; and 0.81-1.00, almost perfect agreement [21]. The study used only anonymized test items and did not involve personal data or human participants. As the experiment was conducted exclusively with LLMs, no additional ethical approval was required.

During the preparation of this manuscript, generative AI (ChatGPT-5.5) was used as an auxiliary tool for language editing (translation, grammar and style corrections, and improvement of readability) and for assistance in developing Python scripts. All AI-assisted code and text modifications were reviewed, verified, and validated by the authors. The authors take full responsibility for the accuracy, integrity, and interpretation of the reported results.

## Results

In the first run, all models demonstrated high performance, answering 80% or more of the test tasks correctly. The highest accuracy was observed for Gemini 3.5 Flash, and the lowest for DeepSeek-V3 (Fig. 1). Pairwise McNemar tests revealed statistically significant differences between Gemini 3.5 Flash and Grok 4 (93.33% and 84.00% respectively, p = 0.0022) and between Gemini 3.5 Flash and DeepSeek-V3 after adjustment for multiple comparisons (93.33% and 80.00% respectively, p = 0.0001).

**Fig. 1.**
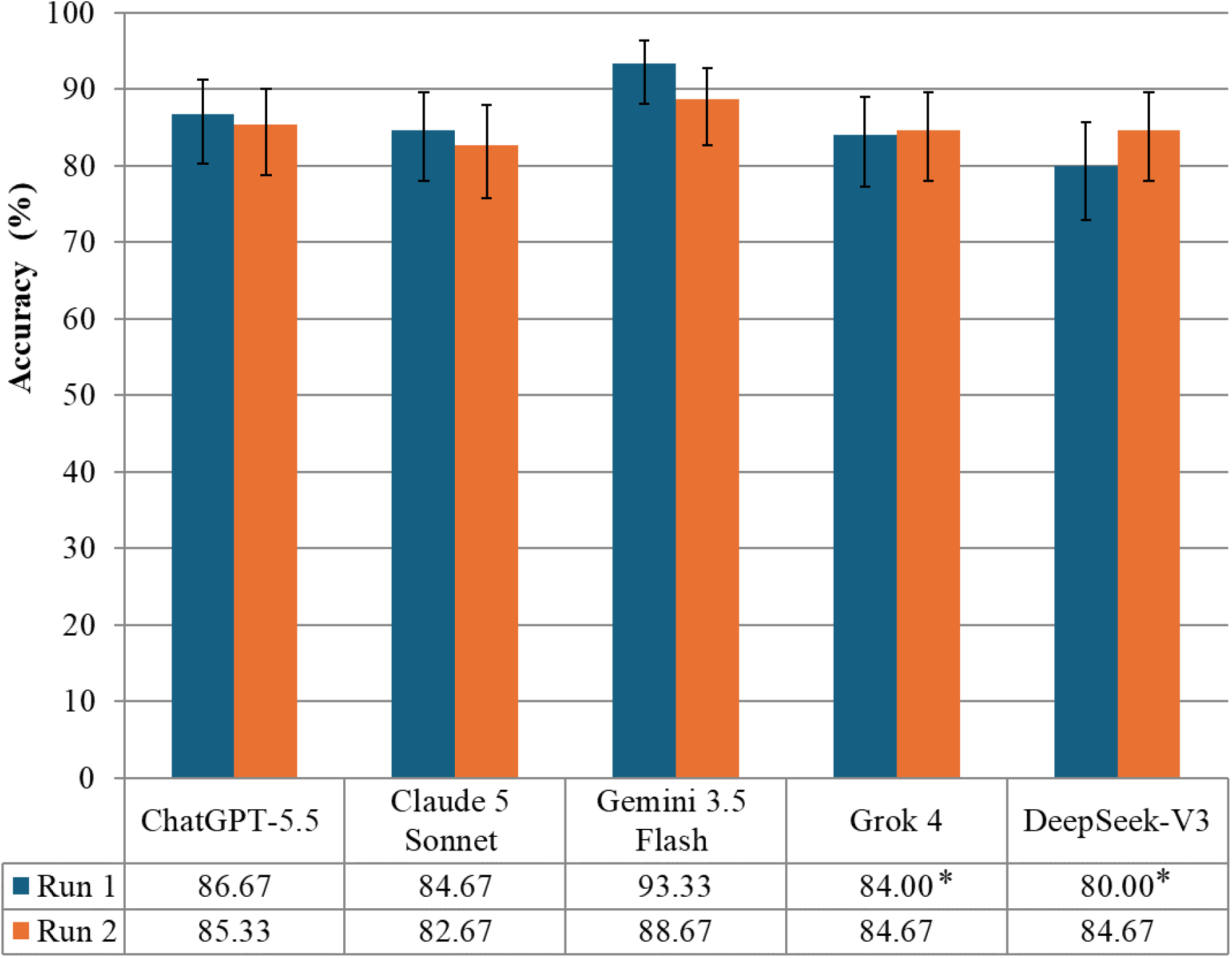
Performance of Large Language Models Across Two Runs, %. Notes. * - the difference is statistically significant, compared to Gemini 3.5 Flash. Error bars indicate 95% confidence intervals.

In the second run, all models showed an accuracy of over 80%. The accuracy of the ChatGPT-5.5, Claude 5 Sonnet and Gemini 3.5 Flash models decreased compared to the first run, while the accuracy of the Grok 4 and DeepSeek-V3 increased. Gemini 3.5 Flash demonstrated the largest observed decrease in accuracy (-4.66%), and the largest increase was for DeepSeek-V3 (+4.67%). However, no statistically significant differences were found in this run as well as between the first and second runs for any of the models. These changes may reflect stochastic variability rather than systematic performance differences.

All models showed R2R response consistency of 90% and above across two independent runs (Table 1).

**Table 1.**
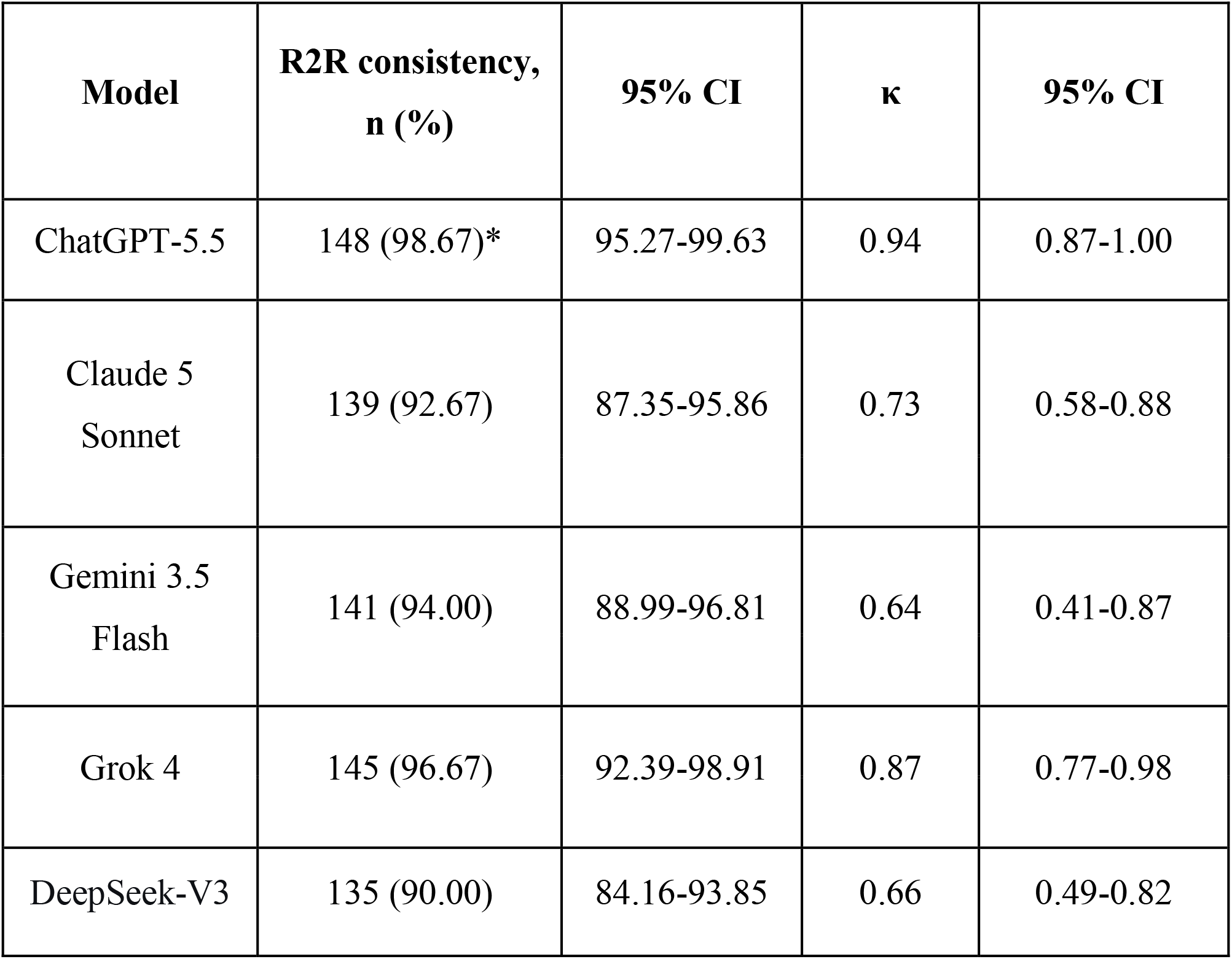
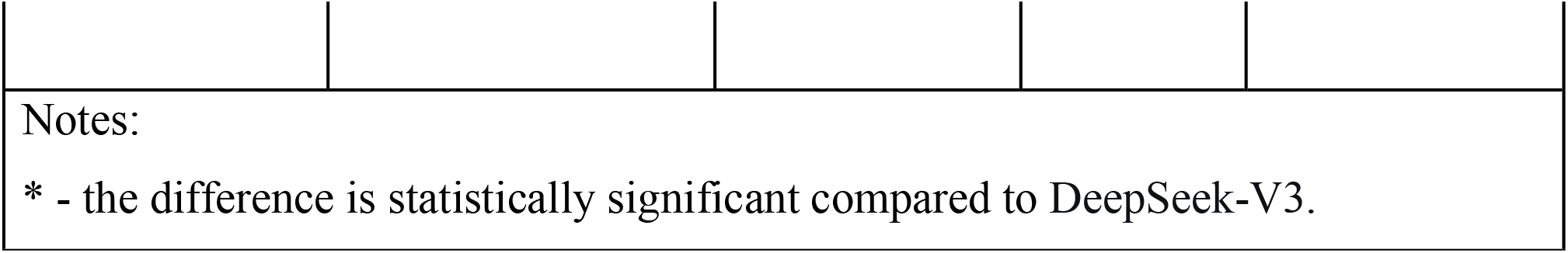
R2R Response Consistency and Agreement Across LLMs in Two Independent Runs.

| **Model** | **R2R consistency,**<br>**n (%)** | **95% CI** | **κ** | **95% CI** |
| --- | --- | --- | --- | --- |
| ChatGPT-5.5 | 148 (98.67)* | 95.27-99.63 | 0.94 | 0.87-1.00 |
| Claude 5<br>Sonnet | 139 (92.67) | 87.35-95.86 | 0.73 | 0.58-0.88 |
| Gemini 3.5<br>Flash | 141 (94.00) | 88.99-96.81 | 0.64 | 0.41-0.87 |
| Grok 4 | 145 (96.67) | 92.39-98.91 | 0.87 | 0.77-0.98 |
| DeepSeek-V3 | 135 (90.00) | 84.16-93.85 | 0.66 | 0.49-0.82 |

Notes:
\* - the difference is statistically significant compared to DeepSeek-V3.

The highest one was observed for ChatGPT-5.5 and the lowest for DeepSeek-V3 (98.67% and 90.00% respectively). The difference in R2R consistency was statistically significant only between these two models (p = 0.0009). At the same time, models agreement, assessed based on the κ, ranged from 0.64 for Gemini 3.5 Flash to 0.94 for ChatGPT-5.5. Thus, Gemini 3.5 Flash, Claude 5 Sonnet and DeepSeek-V3 showed substantial agreement in responses while Grok 4 and ChatGPT-5.5 showed almost perfect agreement.

In terms of models’ responses stability and consistency, Gemini 3.5 Flash showed the highest result, returning the correct answer in both runs for 88.00% of MCQs, whereas the lowest value (77.33%) was recorded for DeepSeek-V3 (Fig. 2). The difference between these two models was statistically significant (p = 0.0008).

**Fig. 2.**
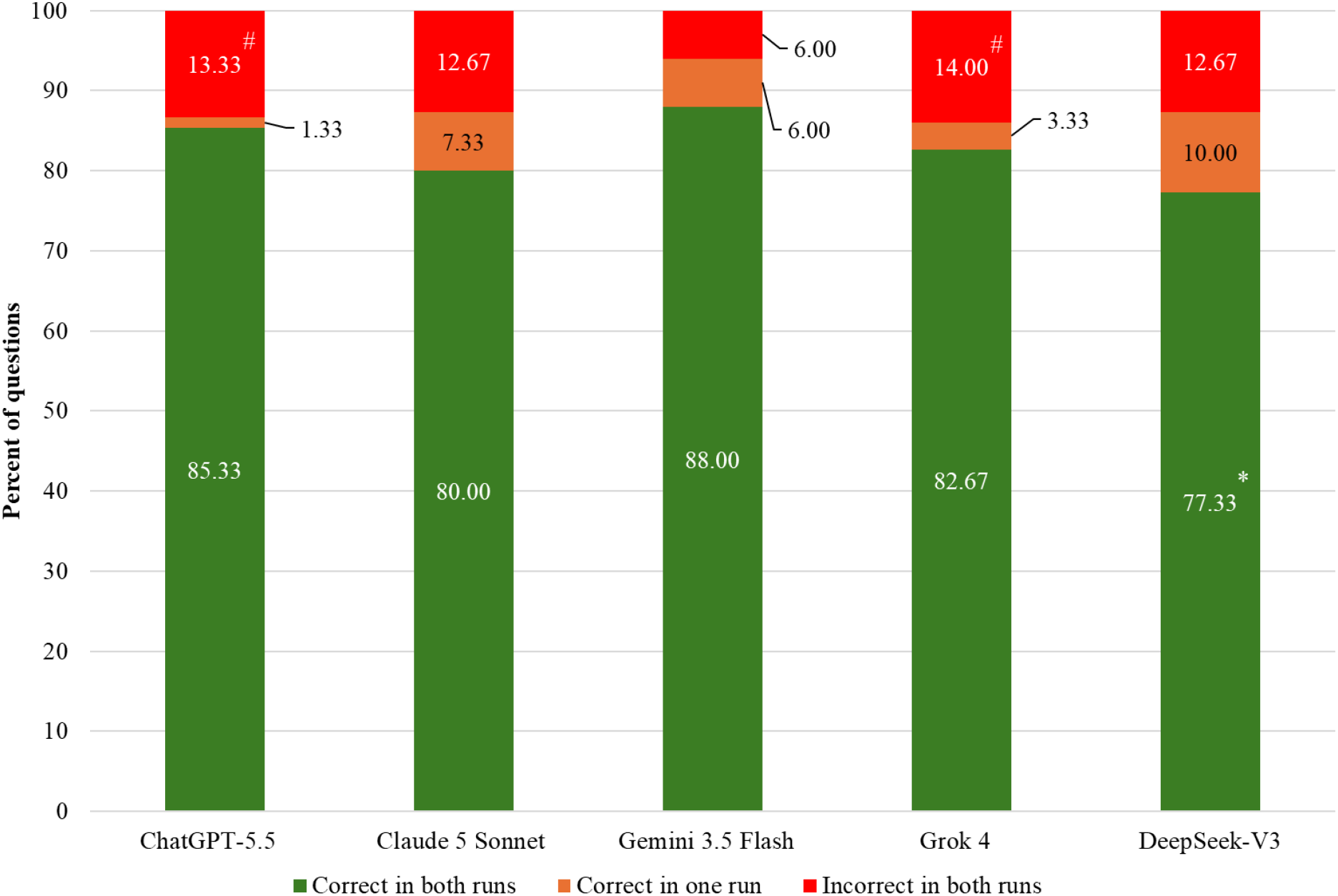
Stability of LLMs’ Response Correctness Across Two Runs (%). Notes: * - the difference is statistically significant, compared to Gemini 3.5 Flash; # - the difference is statistically significant, compared to Gemini 3.5 Flash.

Across both independent runs, Grok 4 demonstrated the highest proportion of incorrect responses (14.00%), whereas Gemini 3.5 Flash showed the lowest (6.00%). The proportion of incorrect responses for Gemini 3.5 Flash was statistically significantly lower compared with ChatGPT-5.5 (13.33%, p = 0.0055) and Grok 4 (14.00%, p = 0.0033).

## Discussion

The results of this study demonstrated that all five evaluated LLMs achieved high accuracy when performing test tasks of the Krok 2 licensing examination in the specialty Physical Therapy. In both independent runs none of the models showed accuracy below 80% and confidently crossed the threshold of ≥ 64%, which corresponds to the criterion for successful completion of this examination when assessing education seekers. Gemini 3.5 Flash demonstrated the highest accuracy (93.33% and 88.67% in the first and second runs, respectively) and achieved the highest overlap of correctly solved test items between the two runs (88.00%). This confirms that modern LLMs are capable of successfully performing not only general medical licensing exams such as the USMLE, but also highly specialized test tasks.

Our results are consistent with the current trend of rapidly increasing effectiveness of LLMs in standardized medical licensing exams. A 2025 systematic review and network meta-analysis of 120 LLM assessments across ten national medical licensing exam systems in nine languages found that most current models already exceed the passing level for such exams, with the most recent models demonstrating accuracy in excess of 90%. It has been found that the performance of models is significantly affected not only by their generation, but also by the type of exam and the testing language [22].

A particular value of our study is its Ukrainian-language context. It has been previously reported that the accuracy of models can vary significantly depending on the language of both the query and the context provided to the model [22]. Despite these expectations, all models in our study demonstrated high accuracy rates on Ukrainian-language test tasks, highlighting the multilingual capabilities of modern LLMs and their potential for use in Ukrainian clinical and educational settings. At the same time, further multilingual studies remain relevant.

An important aspect of our study is the assessment of model reproducibility across independent runs, which has been rarely considered in previous evaluations that have primarily focused on accuracy-related metrics, such as accuracy, precision, recall, or pass rates. In the current study, Gemini 3.5 Flash achieved the highest accuracy, but ChatGPT-5.5 demonstrated superior R2R consistency (98.67%) and agreement (κ = 0.94). Overall, all models demonstrated a high level of response consistency across runs (>90%), indicating stable performance under identical testing conditions. Importantly, the observed differences between models were primarily related to response stability, whereas differences in mean accuracy were less pronounced. These findings highlight that accuracy and reproducibility represent complementary but distinct characteristics of LLM performance.

Our results emphasize that models with similar accuracy may differ substantially in response stability, suggesting that reproducibility should be considered alongside accuracy when evaluating LLM performance, particularly in medical education and clinical applications. These findings are consistent with recent research indicating that response repeatability and stability are important characteristics for the safe implementation of LLMs in healthcare settings [23,24].

The observed differences between models’ accuracy, R2R response consistency and agreement may result from the combined effect of several interacting factors.

First of all, the described phenomenon may be a manifestation of the kappa paradox when a very high percentage of absolute agreement between raters is accompanied by a surprisingly low value of κ. This is because Cohen’s Kappa takes into account the agreement expected by chance, which is artificially inflated when the data categories are highly skewed, for example, one category or outcome appears in the data set much more often than others [25].

In addition, the studied models differ in architectural solutions, the size and composition of the training corpus, as well as approaches to pre-training, which can differently affect both the ability to correctly answer test items and the stability in repeated runs [26].

Another possible explanation is the probabilistic nature of LLM generation, where variability may arise from decoding algorithms, sampling parameters, and internal generation mechanisms. Consequently, models with similar accuracy may demonstrate different levels of response reproducibility [24]. High accuracy reflects the ability of the model to correctly solve test tasks, while high reproducibility characterizes the reliability of its behavior when reused. This approach corresponds to modern trends in the evaluation of medical LLMs, where more and more attention is paid not only to performance indicators, but also to the repeatability, robustness and reliability of models [27].

Our results indicate that modern LLMs have reached a level of performance that makes them promising tools for supporting physical therapists in their practical work and education. These models may support exam preparation (e.g., Krok 2), explanation of complex topics, generation of educational materials, and development of clinical scenarios, while complementing rather than replacing professional expertise. This human-AI collaboration model represents a promising approach for medical education and clinical practice [28]. Modern review studies also consider personalization of learning and instant feedback as one of the main advantages of using generative AI in medical education [29].

However, the results of our study do not indicate the possibility of using LLMs as an autonomous tool for clinical decision-making. The most reasonable approach seems to be to use LLMs as a kind of “second expert”, which helps to check one’s own reasoning, suggest alternative options for clinical analysis or draw attention to potentially missed aspects of the problem. The final decision should remain with a qualified specialist [30].

### Limitations

When interpreting the results, a number of limitations of the study should be taken into account. First, the assessment was conducted only on the standardized Ukrainian integrated test examination Krok 2 in the specialty Physical Therapy, so the results obtained cannot be directly extrapolated to other medical specialties, international licensing exams or real clinical scenarios.

Second, the analysis included only MCQs test tasks, which do not allow for a full assessment of the models’ clinical reasoning, ability to form differential diagnostic hypotheses, justify decisions or work with open-ended tasks.

The study also did not include multimodal tasks: all questions were text-based and did not contain medical images, so the results obtained do not characterize the models’ ability to analyze visual information.

The stability of the models was assessed based on two independent runs, which is sufficient for a preliminary assessment of repeatability, but a larger number of repetitions would provide a more accurate analysis of variability. An additional limitation is the use of publicly available web versions of the models, which did not allow for standardization of generation parameters (temperature, top-p, seed) that could influence the results. Finally, LLMs are constantly updated by developers, so their performance can change even without changing the model’s name. Accordingly, the results obtained reflect the state of the models only at the time of the study, which emphasizes the need for their regular re-evaluation.

### Prospects for further research

The results highlight several directions for future research on LLM applications in physical therapy and medical education. These include regular evaluation of new model generations, assessment of multimodal LLMs capable of analyzing medical images and movement patterns, and studies based on clinical cases and open-ended tasks to evaluate reasoning abilities. Further research should also investigate the impact of prompt engineering, compare commercial and open-source models, and explore the relationship between accuracy, confidence, and response stability to establish more reliable evaluation criteria for the safe implementation of LLMs in clinical and educational settings.

## Conclusions

All evaluated cloud-based LLMs demonstrated high performance on the Ukrainian-language Krok 2 licensing examination in the specialty of Physical Therapy, with accuracy exceeding the official passing threshold in both independent runs. Gemini 3.5 Flash achieved the highest accuracy and the greatest stability of correct responses across runs, whereas ChatGPT-5.5 demonstrated the highest run-to-run response consistency and agreement. These findings indicate that, although accuracy and response stability may be interrelated, they reflect distinct dimensions of LLM performance and should therefore be assessed jointly when evaluating models for medical education and healthcare-related applications. The consistently high performance observed across all evaluated models supports the potential of contemporary LLMs as auxiliary tool for physical therapy professional practice and education. Further studies should evaluate models performance in other medical specialties, clinical reasoning tasks, multimodal assessments, and larger numbers of repeated runs to better characterize the robustness and reproducibility of LLMs.

## Data Availability

All data produced in the present study are available upon reasonable request to the authors

## Information about the authors

Mykhalko Yaroslav Omelyanovych – Candidate of Medical Sciences, Associate Professor, Associate Professor at the Department of internal and family medicine with the courses of instrumental diagnostics, Uzhhorod National University, Narodna Square, 3, Uzhhorod, Ukraine 88000.

Filak Yaroslav Felixovych - Candidate of Sciences in Physical Education and Sports, Associate Professor, Associate Professor at the Department of Physical Therapy, Rehabilitation, Special and Inclusive Education Uzhhorod National University, Narodna Square, 3, Uzhhorod, Ukraine 88000.

Sabadosh Mariana Volodymyrivna - Candidate of Sciences in Physical Education and Sports, Associate Professor, Associate Professor at the Department of Physical Therapy, Rehabilitation, Special and Inclusive Education Uzhhorod National University, Narodna Square, 3, Uzhhorod, Ukraine 88000

